# Correspondence: Recurrent ECSIT mutation encoding V140A triggers hyperinflammation and promotes hemophagocytic syndrome in extranodal NK/T cell lymphoma

**DOI:** 10.1101/2024.09.19.24314011

**Authors:** Jing Quan Lim, Dachuan Huang, Choon Kiat Ong

## Abstract

Recurrent somatic mutation in the *ECSIT* gene encoding p.V140A was reported in 19.3% (17/88) extranodal natural-killer/T cell lymphoma (ENKTL) and was associated with hemophagocytic syndrome (HPS). However, another cohort of similar geographical descent also had the ECSIT^V140A^ mutation as germline in 20.0% (5/25) of ENKTL and none were somatic. The reanalysis of published data revealed that the reported somatic ECSIT^V140A^ could be germline as well. First-digit analysis on the IDs of sequencing read also found irregularity among the data that initially reported the somatic ECSIT^V140A^ mutation. As such, this study questions the only somatic genetic link of HPS in ENKTL and also introduces a simple well-known algorithm to detect data irregularity in voluminous genomic sequencing data.

## Introduction

Haijun Wen *et al*. reported somatic hotspot mutations (c.419T>C) in the evolutionarily conserved signalling intermediate in Toll pathway (*ECSIT*) gene, encoding ECSIT^V140A^, in 19.3% (17/88) of extranodal natural-killer/T cell lymphoma (ENKTL).^1^ ECSIT^V140A^ was found to associate with the activation of NF-kB, poor prognosis and hemophagocytic syndrome (HPS) by triggering hyperinflammation in ENKTL. However, downloaded data from earlier study by Jiang *et al*. from 2015 revealed that ECSIT^V140A^ is common and occurred in 20% (5/25) of patients with ENKTL with paired whole-exome sequencing (WES) data.^2^

Next, we surveyed the literature for whole-genome sequencing (WGS) or WES of ENKTL, but found none to report somatic ECSIT^V140A^ in ENKTL.^3-6^ The Human Genome Diversity Project also reported this as a polymorphic variant with allele frequency of 15% in the Northern Hans within the East Asian population^7^. As such, we suspected the hotspot mutation could be germline instead and proceeded to reanalyse the recurrent ECSIT^V140A^ mutation that was reported in Haijun Wen *et al*..

### ECSIT^V140A^ somatic mutations in Wen *et al*. could potentially be germline mutations

The original paper, Haijun Wen *et al*., had a discovery cohort of five tumor-normal pairs of WES data and wrote that the c.T419C (ECSIT^V140A^) mutation was first detected in one sample (Sample: Appeared as “NKT1” in online repository for the WES data and “P1” in original paper). We reanalysed these five pairs of tumor-normal WES data. Despite good mappability of the sequenced data (∼99.6%, Table 1), we were not able to detect somatic *ECSIT*:c.T419C mutation according to the methodologies that were described in the original paper and by manual eyeballing of the aligned sequenced data.

**Table 1.** Sequencing statistics, alignment statistics, paired tumor-normal Jaccard similarity coefficients of SNPs and coverage of variant/reference statistics at *ECSIT*:c.T419 of the 10 whole-exome sequencing (WES) samples published in Wen *et al.*.

| Sample_ID | number of reads | number of mapped reads | Mapability of sequenced reads | number of duplicated reads (flagged) | number of mapped bases | Jaccard coefficient with sequenced matched-normal (SNPs) | *ECSIT*:c.T419C (+/- reads) | *ECSIT*:c.T419T (+/- reads) | VAF of *ECSIT*:c.T419 C | Peak coverage at *ECSIT* (NM_16581 exon 3) |
| --- | --- | --- | --- | --- | --- | --- | --- | --- | --- | --- |
| TNKT1 | 21,588,015 | 21,543,500 | 99.79% | 308,012 | 2,060,234,242 | 0.676 | 150/0 | 137/0 | 52.26% | 291x |
| TNKT2 | 35,027,481 | 34,170,796 | 97.55% | 1,728,051 | 2,912,908,917 | 0.856 | 0/0 | 7/0 | 0.00% | 25x |
| TNKT3 | 45,860,150 | 45,593,322 | 99.42% | 2,024,976 | 3,909,704,652 | 0.879 | 1/0 | 6/1 | 12.50% | 18x |
| TNKT4 | 43,108,085 | 43,046,631 | 99.86% | 1,317,556 | 3,747,308,450 | 0.866 | 0/0 | 18/0 | 0.00% | 24x |
| TNKT5 | 42,527,467 | 42,482,584 | 99.89% | 1,544,487 | 3,676,514,037 | 0.883 | 0/0 | 5/0 | 0.00% | 21x |
| NNKT1 | 30,101,599 | 30,048,852 | 99.82% | 568,541 | 2,877,430,460 | - | 5/0 | 46/0 | 9.80% | 51x |
| NNKT2 | 35,013,503 | 34,979,380 | 99.90% | 1,474,004 | 3,009,683,864 | - | 0/0 | 13/0 | 0.00% | 41x |
| NNKT3 | 42,422,156 | 42,363,816 | 99.86% | 2,784,612 | 3,553,892,398 | - | 2/0 | 13/6 | 2/21 | 24x |
| NNKT4 | 35,922,786 | 35,877,101 | 99.87% | 1,912,516 | 3,050,206,731 | - | 0/0 | 19/0 | 0.00% | 26x |
| NNKT5 | 45,297,165 | 45,238,072 | 99.87% | 1,999,056 | 3,883,061,179 | - | 0/0 | 19/1 | 0.00% | 20x |

Footnote: Tumoral and normal sample IDs are prefixed with “T” and “N”, respectively. SNP: single nucleotide polymorphism. VAF: variant allele frequency.

Here, we used Integrative Genomics Viewer^8^ to visualize the aligned data (mapping quality > 0) at the genomic location of *ECSIT*:c.T419 and found that most of the alignments were in ‘forward’ orientation (Table 1: column “Forward-strand alignments”) and not equally distributed with the ‘reverse’ orientation (Table 1: column “Reverse-strand alignments”). This contradicted the paired-end (forward-reverse) read-format from Illumina Genome Analyzer IIx or HiSeq 2000 platform instead of the forward-forward format as mentioned by the original authors. Next, the variant allele frequencies (VAF) of *ECSIT*:c.T419C to be 52% (150/288) and 10% (5/51) in the tumor and matching-normal data of NKT1, respectively. These two observations violated rules (ii) and (iii) in the Online Methods of the original paper that were used for the detection of somatic mutations from the sequencing data. Additionally, *ECSIT*:c.T419C was annotated as “rs145036301” in dbSNP135, with submission ID “ss465480684”, and released publicly on 17^th^ September 2011.^9^ Hence, rule (iv) of the Online Methods that was based on dbSNP132, released in July 2010, was not up-to-date to filter away this potential germline mutation.

Furthermore, we also found sequencing reads supporting *ECSIT*:c.T419C in both tumoral and matching-normal WES data of another sample, NKT3, too (alternate variant/total depth of tumor=1/8x; normal=2/21x) (Table 1). In addition, the Jaccard similarity coefficient^10^ between called sets of single nucleotide polymorphisms from paired tumoral and normal samples of NKT1 (Table 1; 0.676) as compared to the remaining four paired samples (Table 1; range: 0.86-0.88) also suggested that the samples of NKT1 might be mismatched, which might have caused the germline mutation to leak through the somatic variant-calling pipeline.

Hence, our reanalyses suggest that the *ECSIT*:c.T419C mutations in Wen *et al*. could be germline mutations.

### Inconsistent sequencing coverage depths within sequencing data of NKT1 (the first ECSIT^V140A^ ENKTL sample)

We also noticed that the sequencing depth of the genomic locus at *ECSIT*:c.T419 of the tumoral sample of NKT1 was at least ∼12.1-times higher (Table 1, column: “Peak sequencing coverage at *ECSIT* (NM_16581 exon 3)”) than the other four tumoral samples. However, the sequencing throughput for the 10 published samples were relatively similar (Table 1, column: “number of mapped bases”) and were within ∼1.9-times of each other.

Furthermore, the aligned data from these five pairs of samples at genomic loci of frequently mutated genes in ENKTL, such as, *DDX3X, CD274, STAT3* and *JAK3*, were showing relatively similar coverage depths except at the *ECSIT* gene (Supplementary Table 1). These observations suggested that the sequencing data at the genomic locus of *ECSIT* had been further enriched, by an undisclosed technique in the original paper, for sample NKT1 but not for the other samples. Next, we analysed the raw sequencing data further to explain this irregularity of coverages within the samples.

### Data irregularity of NKT1 in Wen *et al*. opposes Benford’s first-digit law

During genomic sequencing runs, different parts of the genome are sequenced randomly and the sequencing reads’ IDs (read_IDs) of aligned sequencing reads from a genomic locus, such as a gene, will present itself to obey Benford’s first-digit law. This holds true if the read_IDs have been assigned in a monotonous increasing order of integers starting from 1 to the voluminous data that is being generated during a sequencing run. As such, first-digit counting of the aligned read_IDs will usually yield the highest count for ‘1’ as compared to the remaining eight digits from ‘2’ to ‘9’.^11,12^

For the basis of understanding how the first-digits of read_IDs from aligned sequencing data will present itself, we first simulated read_IDs for WES data at 30x, 50x and 100x coverages and presented the first-digit histogram of read_IDs in Figure 1a. Next, we also studied how the read_IDs presented themselves to Benford’s first-digit law over lower-quartile (1991 bp), median (2938 bp) and upper-quartile (4486 bp) transcript lengths, with references from National Center for Biotechnology Information Gene database entries^13^, over simulated 30x (Figure 1b), 50x (Figure 1c) and 100x (Figure 1d) coverage depths. Without loss of generality, the first-digit counting of the extracted read_IDs from smaller genomic regions, represented by various transcript lengths, presented themselves very similarly among themselves.

**Figure 1.**
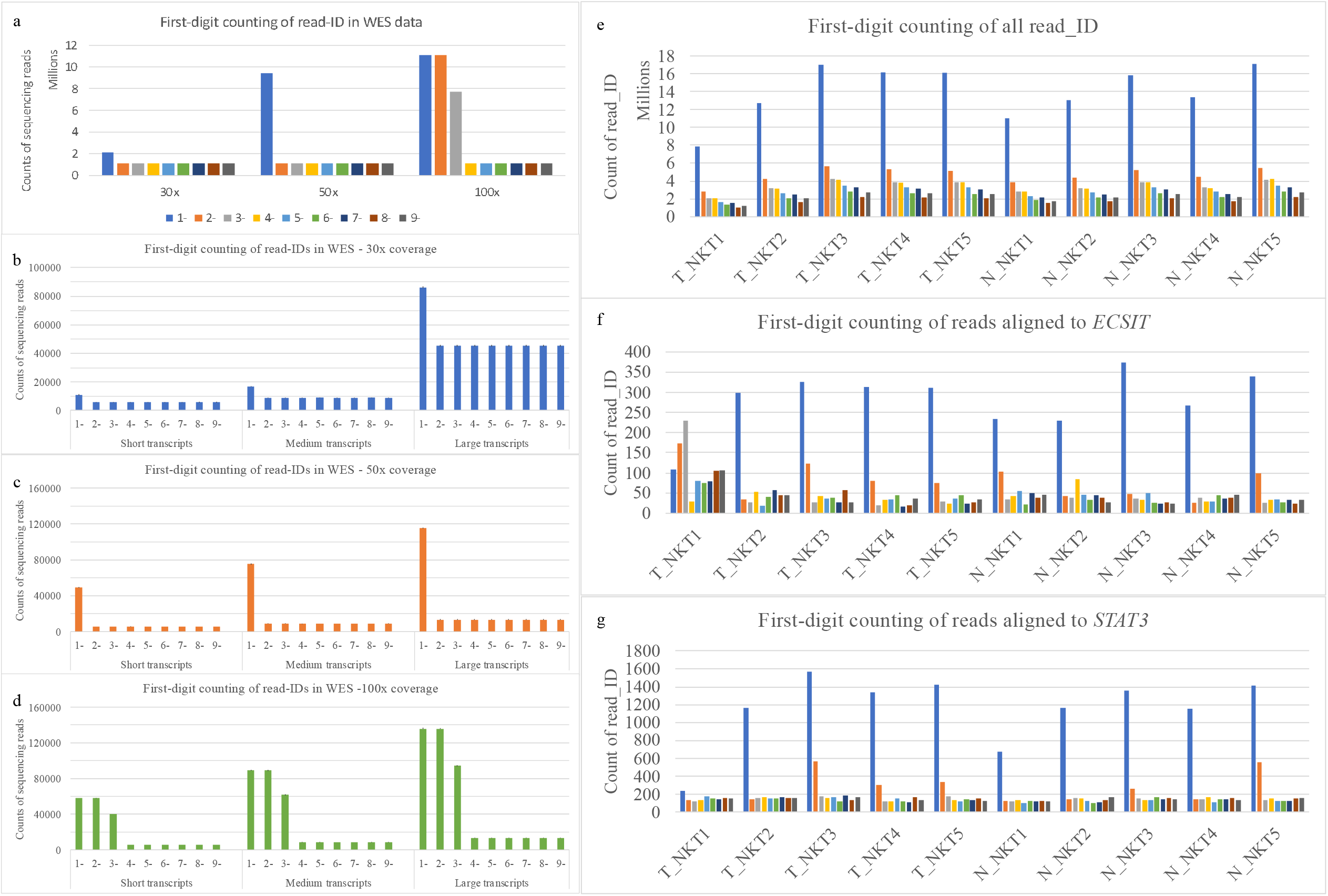
First-digit analysis on the read_IDs of whole-exome sequencing (WES) data. Panel a) Histogram for the counting of first-digit of the read_IDs of *in-silico* WES data of various coverages. Panel b-d) Histogram for the counting of first-digit of the read_IDs at b) 30x c) 50x d) 100x coverage across three different transcript lengths. Panel e) Histogram for the counting of first-digit of the read_IDs of real-world WES data from the initial discovery cohort of Haijun Wen *et al*.. Panel f-g) Histogram for the counting of first-digit of the read_IDs that aligned to f) *ECSIT* and g) *STAT3* gene. Tumoral and normal sample names are prefixed with “T_” and “N_”, respectively.

We then repeated the same analysis on the read_IDs in the five pairs of WES from the original paper (Figure 1e). The first-digit counting of read_IDs from the locus of the *ECSIT* (Figure 1f) was statistically different with the first-digit counts of read_IDs from the loci of frequently reported mutated genes of ENKTL, such as, *DDX3X* (Figure 1g), *CD274, STAT3* and *JAK3* (Supplementary Figure 1), of the tumoral data from NKT1. This analysis confirmed that the WES tumoral data of NKT1 has not been uniformly captured and sequenced, and could have potentially biased the reported outcome.

## Conclusion

Haijun Wen *et al*. reported somatic *ECSIT*:c.T419C in NKT1 within the initial discovery cohort of the five paired tumor-normal WES data. To confirm the somatic-ness of the mutation, we manually inspected the alignments at multiple genomic loci of the WES data and discovered that the mutations were potentially germline. In addition, the aligned read-depth was unexplainably high for the locus of *ECSIT*:c.T419C in the tumoral data of NKT1. We then used first-digit numerical analysis on the read_IDs of the sequencing data and reconfirmed that this subset of data was irregular with respect to all the remaining samples that formed the initial discovery cohort of the original paper. We suspected there was a spike-in of *in silico* data, but we were unable to pinpoint the exact reason to this observed irregularity within the data of the tumoral sample of NKT1 at the *ECSIT*:c.T419C locus. Given that numerous sequencing studies of close geoepidemiology (East and South Asians) to the original paper have not detected somatic *ECSIT*:c.T419C within their data, we hope that the original authors can provide explanations to our observations of the irregularity on their data and if *ECSIT*:c.T419C should still be considered as somatic for future studies.

## Data availability

The WES data published by the original paper was downloaded from NCBI’s repository (BioProject ID: PRJNA422020). The full first-digit counting of read_IDs from 100 runs of simulated WES data over an assumed 55 Mbp exome with 2×75 bp reads is recorded in Supplementary Table 2.

## Declarations of conflict

The authors declared no conflict of interest.

## Authors contributions

JQL conceived the study, analyzed the data, wrote the manuscript and coordinated the study. DCH and CKO revised and approved the manuscript.

## Inclusion & ethics statement

The roles and responsibilities of all listed authors were agreed upon throughout the study and have fulfilled the criteria for authorship required by Nature Portfolio journals. This study was not restricted in any settings of the researchers, and have minimized potential stigmatization, incrimination, discrimination to all its readers.

## Notes

### Competing Interest Statement

The authors have declared no competing interest.

### Funding Statement

JQ Lim was supported by MOH-OFYIRG22jul-0017. The funder did not take part in the design and analyses of the study.

### Author Declarations

The study used ONLY openly available human data that were originally located at BioProject ID: PRJNA422020. An additional dataset from Jiang et al, Nature Genetics 2015 was downloaded from SRP057085. All other data are simulated data.

