## Supplementary Table 1 for "Correspondence: Recurrent ECSIT mutation encoding V140A triggers hyperinflammation and promotes hemophagocytic syndrome in extranodal NK/T cell lymphoma"

Matters Arising

Supplementary Table 1. Peak and relative coverages from the whole-exome sequencing data published by the original authors of Wen *et al*. at the *ECSIT*, *CD274*, *DDX3X*, *JAK3* and *STAT3* genes.

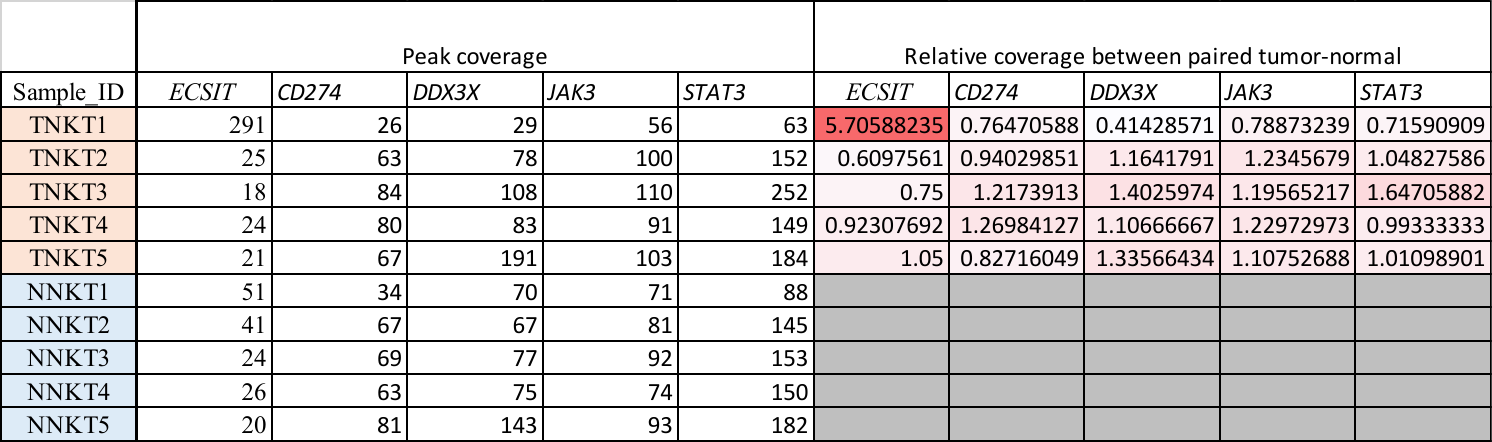
