## Supplementary Figure 1 for "Correspondence: Recurrent ECSIT mutation encoding V140A triggers hyperinflammation and promotes hemophagocytic syndrome in extranodal NK/T cell lymphoma"

Matters Arising

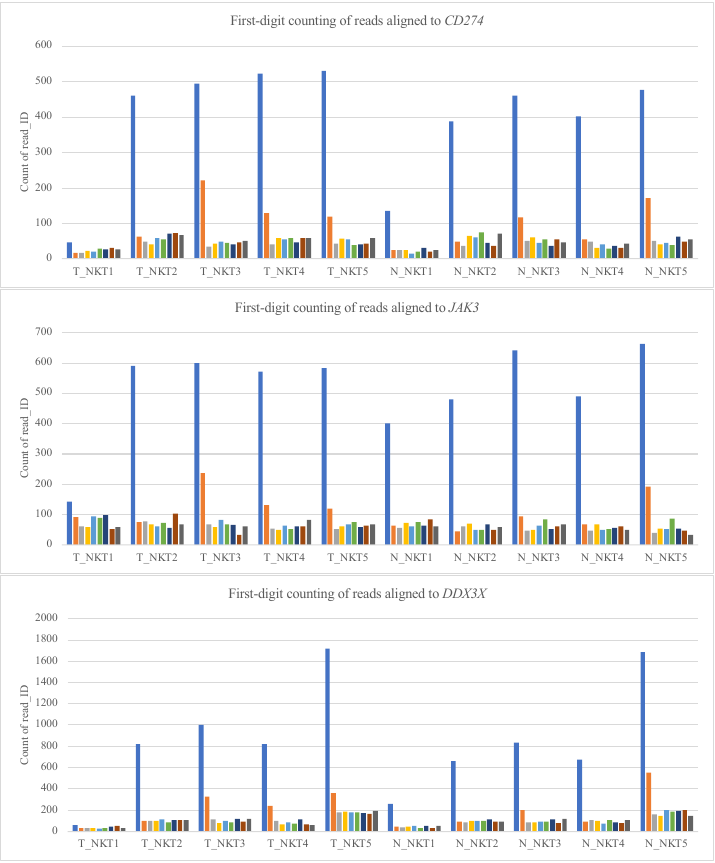

Supplementary Figure 1. Histogram for the counting of first-digit of the read_IDs that aligned to the genomic loci of *CD274*, *JAK3* and *DDX3X*.
